# Aakhyan: An AI-Powered Vernacular Patient Communication Platform for Oncology in Resource-Limited Settings

**DOI:** 10.64898/2026.04.15.26350965

**Authors:** Debaraj Shome Purkayastha

**Affiliations:** Department of Oncosurgery, Silchar Cancer Centre (Assam Cancer Care Foundation), Silchar, Assam, India

## Abstract

Inadequate discharge communication is a well-documented contributor to medication non-adherence, missed follow-ups, and preventable readmissions across healthcare systems worldwide. In resource-limited oncology settings — where patients are often low-literate, speak non-dominant languages, and manage complex multi-drug regimens — this problem is acute and largely unaddressed. We present Aakhyan, a vernacular patient communication platform that addresses the full post-discharge arc: from converting English-language discharge summaries into structured, voice-based vernacular explanations, through medication adherence support, to proactive follow-up management — all delivered via WhatsApp. The architecture is novel in its strict separation of concerns: a vision-language model performs structured JSON extraction from discharge images; all patient-facing content is generated deterministically from clinician-approved templates with community-sensitive vocabulary registers. This design eliminates the hallucination risk inherent in generative AI patient communication (documented at 18–82% in prior studies) while preserving the extraction capability of large language models. The platform supports four language registers — Bengali, Hindi, simplified English for tribal populations, and Assamese — with text-to-speech synthesis across all registers, including a custom grapheme-to-phoneme engine developed for Assamese phonology. Beyond discharge communication, the platform includes scheduled medication nudges, interactive follow-up reminders, and a Daily Availability and Patient Notification System (DAPNS) that notifies patients the evening before their follow-up whether their doctor and required investigations are available — preventing wasted trips by rural patients who travel 2–6 hours to reach the centre. Additional modules are under development to extend the platform toward comprehensive patient care communication. A 100-patient stratified randomised controlled study is planned at Silchar Cancer Centre, with structured teach-back assessment at 48–72 hours post-discharge as the primary comprehension outcome and preliminary clinical efficacy as a secondary objective. This paper describes the clinical rationale, technical architecture, safety framework, and positioning of Aakhyan within the existing literature on mHealth patient communication interventions.

## 1 The Problem: Discharge Communication in Oncology

### 1.1 The Universal Comprehension Gap

Hospital discharge is one of the most information-dense and consequential moments in a patient’s care journey, yet it is consistently one of the most poorly executed. A large observational study of emergency department discharge practices found that at least 78% of patients exhibited comprehension deficits in at least one area of their discharge care, with medication understanding being the lowest-scoring domain at just 41% [1]. Assessment of patient comprehension occurred during only 53% of discharges, and fewer than half of patients were given the opportunity to ask follow-up questions.

Patient recall degrades rapidly. A University of Michigan study found that while more than 90% of patients expressed confidence in their recollection 24–48 hours post-discharge, roughly half were inaccurate [2]. A meta-analysis of discharge instruction delivery modalities showed pooled correct recall rates of 47% for verbal-only information, 58% for written, and 67% for video-based delivery [3]. A systematic review and meta-analysis published in JAMA Network Open demonstrated that communication interventions at discharge significantly reduced readmission rates (9.1% vs 13.5%, RR 0.69), improved medication adherence (86.1% vs 79.0%, RR 1.24), and increased patient satisfaction (60.9% vs 49.5%, RR 1.41) [4].

### 1.2 Oncology-Specific Complexity

Oncology patients face particular challenges. Discharge regimens frequently involve 8–15 medications with varying schedules, food relations, taper protocols, and SOS conditions. Patients commonly misunderstand medication instructions during the transition between inpatient and ambulatory settings, placing them at risk for under- or overdosing [5]. Health literacy has a strong positive correlation with oral chemotherapy adherence (r = 0.707, P < 0.001) [6]. In a tertiary hospital in India, medication errors occurred in 13.6% of observed administrations in oncology, with 81% of adverse drug reactions deemed preventable — inappropriate antiemetic regimens (22%) and lack of supportive care (18%) being leading factors [7].

A survey at one cancer centre found that before implementing a structured discharge education programme, 56% of patients reported not being well-informed about what to expect, 68% were not informed about side effects, and only one patient reported being informed about all available resources [8].

### 1.3 The Indian Context: Where Language, Literacy, and Geography Converge

India’s health literacy landscape compounds these challenges. Less than 40% of the population possesses adequate knowledge about preventive healthcare (National Health Profile, 2022). Only 11% of older adults in rural India demonstrate adequate digital health literacy [9]. A study of 510 residents in a resource-poor rural community in northern India found the lowest scores for “actively managing health” (1.81/5) and “ability to find good health information” (2.65/5) [10].

Northeast India presents a particularly acute version of this problem. The region has the **highest cancer incidence in the country — three times the national average** — with approximately 45,200 new cases reported annually. Assam alone accounts for more than 32,000 cases [11]. Oesophageal cancer, driven by widespread betel nut and tobacco use, occurs at age-adjusted incidences of 19–50 per 100,000 — over ten times the national average [12]. The doctor-to-patient ratio in Assam stands at approximately 1:2,000, double the WHO-recommended minimum of 1:1,000 [13].

Silchar Cancer Centre (Cachar Cancer Hospital and Research Centre), the site of the planned pilot, is a 100-bedded comprehensive cancer centre serving the Barak Valley, Dima Hasao, and neighbouring districts of Tripura, Manipur, and Meghalaya — a catchment area of 30–40 lakh people. Approximately 3,000 new patients (1,700 confirmed cancers) and 15,000 follow-up patients are seen annually. Nearly 80% of patients are daily wage workers, agricultural labourers, and tea garden workers; 75% receive treatment free of charge or at subsidised rates [14].

Cancer treatment abandonment in rural India reaches 62%, driven by travel distance, poor road connectivity, and financial barriers [15]. Loss to follow-up means interrupted chemotherapy cycles, missed surveillance imaging, and delayed detection of recurrence. The problem is not non-compliance — it is non-comprehension compounded by structural barriers.

### 1.4 Language Diversity in the Barak Valley

The Barak Valley has an overwhelming Bengali majority (80.8%, 2011 Census), but it is also home to a diverse linguistic tapestry: Dimasa, Koch Rajbongshi, Meitei, Bishnupriya Manipuri, and various tribal communities including Khasi, Mizo, Hmar, Kuki, and Naga peoples [16]. Bengali is the language of administration and daily life, spoken across a spectrum of regional and community dialects. Hindi serves a migrant working population. English functions as an administrative and educational lingua franca, and is the primary medium through which many tribal patients — conversant in English but not always literate in medical terminology — interact with the healthcare system.

Assamese, the official state language of Assam, is supported with full voice delivery via a commercial TTS provider^1^ paired with a custom rule-based grapheme-to-phoneme (G2P) engine developed specifically for this project. The G2P engine addresses Assamese-specific phonology that commercial TTS models — typically trained on Bengali data — do not handle natively: Assamese fricatives, verb-ending vowel patterns, and the correct pronunciation of Assamese-specific characters. While a small number of mispronunciations remain (notably, certain Assamese words are spoken with Bengali intonation patterns by the underlying model), the output has been validated as intelligible by local listeners. All four active language registers provide voice + text WhatsApp messages.

Discharge summaries at Silchar Cancer Centre are invariably written in English by clinicians. The patient population receives them as incomprehensible documents written in a language and register that, even when technically understood, does not convey actionable meaning about medications, schedules, or danger signs.

## 2 Prior Art and the Gap Aakhyan Fills

### 2.1 mHealth Discharge Communication

The evidence base for mobile health interventions in post-discharge communication is growing. A systematic review of mHealth and teach-back communication examined 17 studies and found that 7 of 11 mHealth studies demonstrated significant hospital readmission reduction, concluding that such tools “can be utilised in resource-constrained settings, especially low- and middle-income countries” [17]. WhatsApp-based interventions specifically have shown promise: a systematic review of WeChat/WhatsApp use among oncology patients (20 studies, 3,110 participants) found statistically significant improvements in pain, medication adherence, self-efficacy, quality of life, and depression [18]. An RCT with 403 diabetes/hypertension patients in Brazil demonstrated a clinically significant 15% increase in medication adherence with WhatsApp educational intervention [19].

However, these interventions share common limitations. They rely on **manually authored** content, require healthcare workers to compose and send messages individually, and offer no mechanism for structured extraction from existing clinical documents. None addresses the fundamental pipeline problem: converting a clinician’s discharge summary into a patient-comprehensible, voice-first, language-appropriate explanation without human authoring at every step.

### 2.2 Generative AI for Patient Communication

Recent studies have explored using large language models (LLMs) to simplify discharge summaries for patients. The results are cautionary:

- A JAMA Network Open study (2024) tested GPT to transform 50 discharge summaries into patient-friendly formats. While understandability improved, **18% of reviews noted safety concerns** including omissions and inaccuracies, with 6% exhibiting hallucinations and 3% containing fabricated medications [20].
- An evaluation in npj Digital Medicine (2024) found “potentially harmful safety issues attributable to the AI tool in 18%” of AI-generated patient discharge instructions [21].
- A systematic framework for assessing LLM hallucination in medical text summarisation (2025) found hallucination rates ranging from **50% to 82%** across models, with prompt-based mitigation only reducing rates from 66% to 44% [22].
- A comparative study found that a template-based approach (SmartGuideline) produced **zero major hallucinations**, whereas GPT-4 generated 6 major hallucinations defined as responses that could “plausibly mislead clinical decision-making” [23].

These findings establish a clear principle: **generative AI cannot be safely used to compose patient-facing medical text**. The hallucination rates are too high, the failure modes are too dangerous, and the mitigation techniques are insufficient. This is the central architectural insight that motivates Aakhyan’s design.

### 2.3 The Unfilled Niche

An exhaustive search of the literature reveals that no existing system combines all of the following capabilities:

| Capability | Closest Prior Art | Aakhyan Innovation |
| --- | --- | --- |
| Vision-based extraction from discharge images | MIRAGE (2024): fine-tuned models on simulated Indian prescriptions, 82% accuracy [24] | Dual-pass extraction from real handwritten/printed oncology discharge summaries with field-level diff |
| Patient-facing text generation | GPT-4, Claude used in prior studies | AI performs extraction only; deterministic templates generate all patient text — zero hallucination by design |
| Community-sensitive registers | Language-concordant instructions exist [25] | Multiple vocabulary registers within the same language (sociolinguistic, not merely linguistic) |
| Voice-first delivery in Indic languages | Standalone Indic TTS APIs | Integrated end-to-end pipeline with custom Assamese G2P: extract > template > TTS > WhatsApp delivery |
| Interactive patient engagement | WhatsApp chatbots exist | Structured keyword menu derived from extracted clinical data (medications, follow-ups, diet) |
| Risk-stratified medication nudges | Generic SMS reminders exist | Nudge protocol calibrated to clinical risk (opioid regimens, custom schedules) |
| Follow-up reminders with verification gate | Appointment reminder systems exist | Reminders fire only after clinician verification of extracted dates — prevents wrong-date reminders |
| Clinician approval gate with audit trail | Human-in-the-loop is recommended [26] | Mandatory approval with tabbed original-image/blueprint comparison, field-level edit tracking |
| Pre-visit availability notification | Appointment reminder systems exist | D-1 evening notification cross-referencing doctor/test availability against patient follow-up schedule — prevents wasted rural trips |

Aakhyan is, to our knowledge, the first platform to implement the complete pipeline from discharge image to voice-based vernacular patient communication — and extend it through the post-discharge follow-up arc — with a deterministic safety architecture throughout.

## 3 System Architecture

### 3.1 Design Philosophy: AI as Classifier, Not Composer

The core safety invariant of Aakhyan is:

**The LLM may only output structured JSON data. All patient-facing content is generated from deterministic, clinician-approved templates. No LLM-written text, advice, or free-form content ever reaches the patient**.

This is not a limitation — it is the primary innovation. The system harnesses the extraction and classification capabilities of vision-language models while eliminating their generative failure modes. The AI acts as a structured data extractor, not a content author.

### 3.2 Six-Layer Pipeline

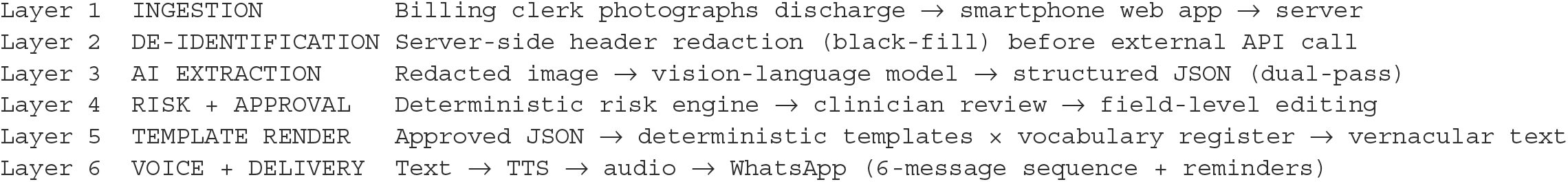

### 3.3 Dual-Pass Extraction with Field-Level Verification

Each discharge image undergoes two independent extractions at temperature 0. The system performs field-level comparison between runs, flagging discrepancies. This is analogous to double data entry in clinical research — a well-established method for ensuring data quality. Fields where the two runs disagree are highlighted on the approval screen for clinician attention.

The extraction prompt covers multiple medication patterns (oral, injection, topical, taper, SOS, continuation, and custom schedules), demographic fields, follow-up events, diet advice, drain status, and safety flags (handwriting detection, strike-through detection, opioid presence).

### 3.4 Community Vocabulary Registers

Aakhyan implements a register system that goes beyond language translation. The architecture supports vocabulary registers that can adapt within a single language to match a patient’s community and regional dialect. While all clinical content — medication instructions, safety warnings, follow-up details — remains identical across registers, everyday words (greetings, food, family) can be adapted to match the vocabulary familiar to a particular community. This signals “this message is for you” through recognisable vocabulary, without altering any clinical meaning.

For the pilot, four registers are active: Bengali (formal), Hindi, simplified English (for tribal patients conversant in English but unfamiliar with medical terminology), and Assamese. The architecture is designed to accommodate additional community-adapted registers — for example, regional Bengali dialects or vocabulary adapted for specific tribal communities — requiring only a vocabulary file and community validation with local speakers. No changes to the template engine, drug lexicon, or delivery pipeline are needed.

This design was informed by the observation that language concordance alone is insufficient. A patient who hears everyday words from their own community — rather than a formal register that feels distant even within the same language — engages with the content differently.

### 3.5 Drug Functional Lexicon and Intelligent Fallback

The system maintains a curated lexicon of medications commonly prescribed at Silchar Cancer Centre, mapping brand names and generic names to functional classes with plain-language explanations in all four registers. For drugs not in the lexicon, a three-tier fallback ensures no medication is silently omitted:

- **Class match**: The AI’s extracted functional class maps to a known category (antibiotic, antiemetic, analgesic, etc.) — the curated class description is used.
- **Helper-drug framing**: For truly novel drugs, the system frames it as a supportive medicine with full scheduling information.
- **Route-specific enrichment**: The framing adapts to oral, injection, topical, or continuation contexts.

The patient always hears: the drug name (matching their tablet strip), what it does in plain language, and when/how to take it. The instruction “take as your doctor directed” — common in discharge summaries and functionally useless — never appears.

### 3.6 Text-to-Speech and WhatsApp Delivery

Rendered vernacular text is synthesised using two TTS providers selected for language quality and Indic language coverage. Audio segments are delivered via WhatsApp as voice messages — the most accessible format for low-literate populations who may struggle with text messages.

| Language | TTS Approach |
| --- | --- |
| Bengali | Commercial multilingual TTS<sup>2</sup> |
| Hindi | Indic-specialised TTS<sup>3</sup> |
| English | Indic-specialised TTS<sup>3</sup> |
| Assamese | Commercial multilingual TTS<sup>2</sup> + custom G2P engine |

The TTS providers were selected after comparative evaluation across naturalness, Indic language coverage, and code-mixing capability (English drug names pronounced naturally within vernacular sentences). For Assamese — a language not natively supported by any production Indic TTS service — a commercial multilingual provider was paired with our custom G2P engine (described below).

#### Custom Assamese G2P engine

No commercial TTS system natively supports Assamese phonology — available models are trained predominantly on Bengali data and systematically mispronounce Assamese-specific sounds. To address this, we developed a custom rule-based grapheme-to-phoneme (G2P) engine that preprocesses Assamese text before TTS synthesis. The engine handles Assamese fricative consonants, verb-ending vowel patterns distinct from Bengali, and the correct rendering of Assamese-specific characters (e.g., ৰ, ৱ). It also applies context-sensitive overrides for medication names — ensuring drug names pass through to the TTS model unmodified while surrounding Assamese text is phonetically corrected. This is, to our knowledge, the first application of a purpose-built Assamese G2P pipeline for healthcare voice delivery.

The Day 0 discharge sequence comprises six messages: introduction, medication explainer (the core 2–3 minute segment), red flags and follow-up information, an interactive command menu, diet advice, and conditional drain care instructions. Patients can reply with single-digit numbers (1–5, 0) at any time to hear their medication schedule, hospital contact, follow-up date, diet advice, or to opt out.

### 3.7 Progressive Web Application

The system is delivered as a Progressive Web Application (PWA) — installable on the billing clerk’s smartphone home screen, providing an app-like experience without requiring distribution through app stores. This is critical for deployment in settings where institutional app distribution is impractical.

### 3.8 English Language Pipeline

A significant innovation is the inclusion of a simplified English register. Northeast India’s tribal populations — Khasi, Mizo, Hmar, Kuki, Naga, and others — often speak English as a lingua franca acquired through missionary education and administrative exposure. They can read and converse in English, but may not parse the dense medical terminology of a discharge summary (“Tab Pantoprazole 40mg 1-0-0 BBF × 14d” conveys little to someone unfamiliar with medical shorthand).

The English register translates medical jargon into plain English:

- “1-0-1 after food” becomes “one tablet in the morning after breakfast, one tablet at night after dinner”
- “BBF” becomes “before breakfast, on an empty stomach”
- “SOS” becomes “only when needed — for example, if you feel nauseous”
- “OPD review” becomes “outpatient clinic visit”

This register uses the same deterministic template architecture as the Bengali and Hindi registers, ensuring no clinical information is lost or hallucinated in the simplification process.

### 3.9 Daily Availability and Patient Notification System (DAPNS)

A significant contributor to treatment abandonment in rural oncology is the wasted trip: a patient or caregiver travels 2–6 hours to reach the cancer centre, only to find that their consultant is on leave or a required investigation (PET-CT, CT scan) is under maintenance. The resulting frustration, financial loss, and erosion of trust in the healthcare system accelerates dropout from treatment protocols.

Aakhyan addresses this with DAPNS — a module that notifies patients the **evening before** their follow-up appointment whether their doctor and required investigations are available. The timing is critical: rural patients in the Barak Valley typically depart home at 4–5 AM. A morning-of notification arrives too late. Each afternoon, a designated staff member confirms the availability of consultants and diagnostic facilities for the following day. The system cross-references this against scheduled follow-ups and sends personalised notifications (text + audio) to affected patients by 7 PM.

A core design principle of DAPNS is that the system **never discourages a patient from attending** their appointment. When a doctor or investigation is unavailable, the notification advises the patient to contact the hospital Information Desk — it informs, but does not make clinical decisions about whether a patient should travel. This distinction is critical in a population where treatment abandonment is already high: the system must reduce wasted trips without inadvertently becoming a barrier to care.

DAPNS is deployed with a phased validation approach, and uses the same template architecture, vocabulary registers, TTS providers, and WhatsApp delivery infrastructure as the discharge pipeline.

## 4 Safety Architecture

### 4.1 Defence in Depth

Aakhyan implements multiple independent safety layers:

- **PII Protection**: Header region of discharge images is redacted (black-fill) before any external API call. Patient identifiers never leave the local server for AI processing. Compliant with the Digital Personal Data Protection Act (DPDPA) 2023 [27].
- **Extraction Verification**: Dual-pass extraction with field-level diff catches inconsistencies. Extraction confidence scoring flags uncertain fields.
- **Risk Classification**: A deterministic policy engine classifies every discharge as GREEN, AMBER, or RED based on medication risk (opioids, anticoagulants, insulin), extraction quality, and clinical complexity. RED discharges require the discharging consultant’s personal approval.
- **Clinician Approval Gate**: No message reaches a patient without explicit clinician approval. The approval screen presents the original discharge image alongside the extracted blueprint in a tabbed interface, forcing visual comparison. Every edit is logged with editor identity and timestamp.
- **Template Determinism**: Patient-facing text comes exclusively from version-controlled deterministic templates with vocabulary variables. The content is auditable, reproducible, and free of generative AI artifacts.
- **Red-Flag Symptoms**: Warning symptoms (fever, bleeding, severe pain, etc.) are hardcoded by treatment type (chemotherapy, surgery, radiation, palliative). They are never extracted or generated by the AI — they are always present, always correct.
- **Follow-Up Verification Gate**: Reminders for follow-up appointments fire only when a clinician has explicitly verified the extracted date. This prevents wrong-date reminders — identified as a top project-killing risk.

### 4.2 Why Not Just Use LLMs to Write Patient Messages?

The temptation to use a generative model end-to-end — extracting data and composing the patient message in one step — is understandable. It would be simpler to build. But the evidence is unambiguous:

- 18% of AI-generated patient discharge instructions contain potentially harmful safety issues [21]
- LLM hallucination rates in medical text summarisation range from 50–82% [22]
- 3% of AI-simplified discharge summaries contained fabricated medications [20]

For a system that will tell a cancer patient how to take their morphine or when to return for chemotherapy, these error rates are unacceptable. Aakhyan’s template architecture reduces the patient-facing hallucination rate to **zero by design** — not by mitigation, but by elimination.

## 5 Pilot Study Design

### 5.1 Study Parameters

A prospective, randomised, controlled, single-centre, open-label preliminary efficacy and feasibility study with stratified randomisation by discharging consultant is planned at Silchar Cancer Centre. 100 patients (50 intervention, 50 control) will be enrolled, with block randomisation using random block sizes (4 and 6) to prevent prediction. The Principal Investigator is Dr. Debaraj Shome Purkayastha (MS, Oncosurgery), with co-investigators Dr. Rahuldeb Midya (DrNB Surgical Oncology) and Dr. Yousuf Farid Choudhury (MS ENT, Fellowship Head & Neck Surgery) responsible for the clinician approval workflow and data verification.

### 5.2 Primary Outcomes

#### Feasibility outcomes

coverage rate (target ≥80%), extraction accuracy (target ≥95% field-level), approval turnaround (target median ≤30 minutes), delivery success rate (target ≥95%), message read rate, patient engagement (target ≥40% keyword reply usage), opt-out rate (alarm threshold >10%).

#### Comprehension outcome (hypothesis-generating)

Structured teach-back assessment at 48–72 hours post-discharge via phone call by a blinded assessor. Five domains (medications, frequency/timing, food relation, red flag symptoms, follow-up date), each scored 0–2, total 0–10. Adequate comprehension threshold: ≥7/10.

### 5.3 Natural Timepoint Separation

The system has multiple components, but measurement timepoints naturally isolate their effects:

| Timepoint | Measures | Component Tested |
| --- | --- | --- |
| 48–72h teach-back | Comprehension of discharge instructions | Discharge explainer + medication nudges |
| First follow-up | Attendance, timeliness, preparedness | Follow-up reminder system |
| 30-day outcomes | ED visits, adherence, satisfaction | Whole system cumulative |
| Keyword usage logs | Which features patients use | Patient-initiated engagement |
| DAPNS availability accuracy | Notification matched actual availability | Daily availability system |

### 5.4 Reporting Standards

The study will be reported per the CONSORT extension for pilot and feasibility trials and the WHO mERA (mHealth Evidence Reporting and Assessment) checklist [28], which specifies 16 items covering infrastructure, technology platform, intervention content, usability testing, data security, and contextual adaptability. Results will be published regardless of outcome (positive, negative, or inconclusive), and the CTRI record will be updated with results within 12 months of study completion. If the intervention proves beneficial, control-arm patients will be offered the Aakhyan service post-study.

#### Anticipated timeline

IEC submission (April 2026) > approval (May 2026) > CTRI registration and dry run (May–June 2026) > enrollment (June–September 2026) > 30-day follow-up complete (October 2026) > manuscript submission (December 2026).

### 5.5 Ethical Considerations

- IEC application submitted to SCC/ACCF Institutional Ethics Committee (16 April 2026). CTRI registration planned within 7 days of IEC approval.
- Written informed consent in Bengali, Hindi, English, or Assamese (thumb impression with independent witness acceptable for illiterate patients; legally authorised representative for incapable patients)
- Minimum 15-minute waiting period between information provision and consent signing; delayed consent option within 24 hours via phone confirmation
- Diagnosis disclosure preference captured as a separate signed consent item before any message content is discussed — the patient chooses whether voice messages mention their specific diagnosis or use a generic term
- Community register system described as “culturally adapted health communication using community-appropriate vocabulary” — not religious or ethnic profiling
- DAPNS consent captured as a separate consent item: “We may send you WhatsApp messages about doctor and test availability on your follow-up days” — future tense, no timeline commitment, ensuring DAPNS can be phased in without re-consenting enrolled patients
- DPDPA 2023 compliance: ephemeral image capture, server-side PII redaction before external API calls, encrypted storage, data processor agreements with all third-party AI and TTS providers. Patients may request data erasure under DPDPA Section 17(2)(b), subject to the research exemption. Data retention: 5 years from study completion.

## 6 Cost-Effectiveness and Scalability

### 6.1 Per-Patient Cost

| Item | Cost (100 patients, 6 months) |
| --- | --- |
| Cloud hosting (Oracle Cloud) | INR 15,000 (INR 2,500/month) |
| Domain + SSL + monitoring | INR 3,000 (INR 500/month) |
| AI extraction — vision-language model (dual-pass)□ | INR 3,000–5,000 (INR 30–50/discharge) |
| TTS — Indic-specialised provider (Hindi/English)<sup>3</sup> | INR 1,000–1,500 (INR 10–15/patient) |
| TTS — multilingual provider (Bengali/Assamese)<sup>2</sup> | INR 2,000–3,500 (INR 20–35/patient) |
| WhatsApp delivery (WAHA Plus, self-hosted) | INR 9,600 (INR 1,600/month) |
| Smartphone + SIM for billing desk | INR 15,000 (one-time) |
| Data SIM (6 months) | INR 2,000 (one-time) |
| **Total** | **INR 40,000–55,000** |
| **Per patient (variable costs)** | **INR 100–150 (~USD 1.20–1.80)** |

The per-patient variable cost (AI extraction + TTS) is comparable to a single basic blood test. For context, a standard CBC at a private laboratory in Silchar costs approximately INR 200–400. The study is entirely self-funded by the Principal Investigator.

### 6.2 Scalability Path

The architecture is designed for straightforward replication. The pipeline is stateless — each discharge is processed independently — and adding a new centre requires only configuring the consultant list, adding any centre-specific drug lexicon entries, and provisioning a WhatsApp session. No retraining of AI models or modifications to the safety architecture are needed. The system can be maintained by a single technically literate staff member at each site.

Post-pilot, migration to the Meta Business API for WhatsApp would enable interactive menus, a verified business badge, and broader messaging capabilities. IVR telephony fallback is planned to address the estimated 30% of patients without WhatsApp access.

## 7 Broader Significance

### 7.1 A Replicable Model for LMIC Oncology

Aakhyan addresses a problem that exists wherever cancer care is delivered to populations with limited health literacy — which is to say, most of the world. The modular, template-based architecture means that adding a new language requires only: creating a vocabulary register (150+ keys) and optionally configuring TTS. Adding a new communication module (such as DAPNS) requires no changes to the core safety framework, language infrastructure, or delivery pipeline. No retraining of AI models, no new medical ontologies.

India’s cancer burden is projected to reach 1.56 million new cases annually (ICMR NCRP, 2024), with the lifetime risk of developing cancer at 11.0% [29]. The concentration of treatment in urban centres and the absence of structured discharge communication in most regional cancer centres creates a systemic gap that technology can address at scale.

### 7.2 The Template-Based Safety Paradigm

Aakhyan’s architectural decision — to use AI exclusively for extraction and classification while generating all patient-facing content from deterministic templates — may prove to be its most transferable contribution. As healthcare systems worldwide explore AI-powered patient communication, the temptation to use generative models end-to-end will be strong. The evidence reviewed in this paper suggests that this approach is premature for safety-critical clinical communication.

The template paradigm offers a middle path: leveraging AI where it excels (extracting structured data from unstructured documents) while maintaining human-authored, clinician-approved, version-controlled content for everything that reaches the patient. This is not a temporary compromise — it is an architectural principle that eliminates an entire category of risk.

### 7.3 Addressing Treatment Abandonment Through Proactive Communication

Section 1.3 noted that cancer treatment abandonment in rural India reaches 62%. While the primary drivers are structural — travel distance, financial barriers, poor road connectivity — a significant contributor is the **wasted trip**: a patient loses a day’s wages, arranges transport, and travels hours to find their consultant on leave or a required investigation unavailable. Each wasted trip compounds the perception that the healthcare system is unreliable and erodes the patient’s (and caregiver’s) willingness to continue treatment.

DAPNS directly addresses this by converting the follow-up reminder from a passive “come tomorrow” to an active “come tomorrow — your doctor is here and your tests are ready” or, critically, “call the Information Desk before you come.” The D-1 evening timing ensures the information reaches the patient before the point of no return (the early-morning departure common in the Barak Valley’s geography). The system never makes a clinical decision about whether a patient should travel — it informs, and the patient decides.

With DAPNS, Aakhyan evolves from a discharge communication tool into a patient communication platform that addresses the full post-discharge arc — from comprehension at discharge, through medication adherence, to follow-up continuity. A patient who understands their discharge instructions but abandons follow-up due to wasted trips still has a poor outcome. Closing both gaps — comprehension and continuity — is the objective, with additional modules under development to extend the platform toward comprehensive patient care communication.

### 7.4 Community-Sensitive Health Communication

The vocabulary register system addresses a dimension of health communication that is largely unexplored in the mHealth literature. Existing multilingual health interventions translate content between languages; Aakhyan adapts within a language to match the patient’s community. This is a recognition that language concordance alone is insufficient — a Bengali-speaking patient from a rural Cachar village and a Bengali-speaking patient from Silchar town may speak the same language but use different everyday words for greetings, food, and family. Similarly, a Dimasa-speaking patient from the Dima Hasao hills, a Mizo-speaking patient from southern Mizoram, and a Bengali-speaking patient from the Barak Valley plains all present to the same cancer centre with fundamentally different linguistic and cultural contexts.

The inclusion of a simplified English register for tribal populations further extends this principle. Many patients from Khasi, Mizo, Hmar, Kuki, and Naga communities are conversant in English but do not need translation — they need demystification. Medical English (“Tab Pantoprazole 40mg 1-0-0 BBF × 14d”) is a specialised sublanguage even for English speakers. The English register translates from medical English to plain English, using the same template infrastructure that renders Bengali and Hindi.

### 7.5 Language Expansion Roadmap

#### Current support (April 2026)

**Current support (April 2026):**
| Language | TTS Approach | Delivery Mode | Status |
| --- | --- | --- | --- |
| Bengali | Commercial multilingual TTS<sup>2</sup> | Voice + Text | Active |
| Hindi | Indic-specialised TTS<sup>3</sup> | Voice + Text | Active |
| English | Indic-specialised TTS<sup>3</sup> | Voice + Text | Active |
| Assamese | Commercial multilingual TTS<sup>2</sup> + custom G2P | Voice + Text | Active |

#### Development methodology

New language registers are developed using machine translation for initial vocabulary drafts, but runtime patient-facing content uses only curated, clinician-reviewed vocabulary files — machine translation output never reaches patients directly. Each register undergoes vocabulary review by native speakers and TTS intelligibility testing with local listeners before activation.

#### Planned expansions (requiring native speaker validation)

- **Northeast Indian languages:** Dimasa, Bodo, Meitei (Manipuri, Meitei Mayek script), and Nagamese — reflecting the tribal and indigenous communities served by Silchar Cancer Centre and neighbouring referral centres in Manipur, Meghalaya, and Nagaland. Each requires native speaker vocabulary validation before deployment.
- **Major Scheduled Languages with TTS support:** Tamil, Telugu, Kannada, Malayalam, Marathi, Gujarati, Odia, and Punjabi are straightforward additions given existing TTS coverage from current providers (11+ Indian languages across Indic-specialised and multilingual TTS services). The bottleneck is vocabulary review and clinical terminology validation, not technology.
- **Self-hosted AI model:** Migration to a self-hosted extraction model is on the long-term roadmap to eliminate cross-border data transfer entirely.

#### Modular design

Adding a new language requires only: (1) creating a vocabulary register (150+ keys), (2) adding functional class descriptions and drug lexicon entries, and (3) optionally configuring TTS if a provider supports that language. The text-only delivery path eliminates TTS availability as a blocker — any language can be supported via text messages at near-zero marginal cost. Voice delivery is additive, not prerequisite.

## 8 Limitations

- Non-blinded intervention (patients know they received messages)
- Teach-back at 48–72 hours partially conflates the discharge explainer with medication nudges (Day 1–3), though follow-up reminders have not yet fired — comprehension measurement primarily reflects the discharge explainer
- WhatsApp-only delivery creates selection bias (~30% of eligible patients excluded)
- Single-centre study at a specialised cancer centre
- Pilot framing with preliminary efficacy assessment — not powered for definitive efficacy
- Cannot fully isolate individual component effects without factorial design; natural timepoint separation provides partial insight
- Language register assignment by billing clerk — while reliable in practice, introduces a judgement-based step
- Principal Investigator is the system developer — mitigated by blinded teach-back assessment, pre-registered outcomes, and independent co-investigators for the approval workflow
- Bengali and Assamese TTS use a commercial multilingual provider rather than an Indic-specialised service (which does not support Assamese). While the selected voice produces intelligible output in both languages, some phonological limitations remain in Assamese due to the underlying model’s Bengali training bias.
- Cross-border data transfer to US/EU servers occurs for AI extraction and Bengali/Assamese TTS synthesis. Data is de-identified before transfer, but this remains a limitation addressed in the long-term roadmap via self-hosted model deployment.
- DAPNS provides previous-day availability information; same-day changes (e.g., consultant called away for emergency surgery overnight) are not covered in the pilot phase. A day-of correction mechanism is planned post-pilot.
- DAPNS relies on hospital staff reporting availability each afternoon — the system’s effectiveness is bounded by this human-dependent data entry step.

## 9 Conclusion

Aakhyan represents a new category of clinical communication platform: one that harnesses the extraction power of modern vision-language models while maintaining absolute determinism in patient-facing content. It addresses the full post-discharge patient journey — from the comprehension gap at discharge, through medication adherence, to follow-up continuity — with an architecture that is safe by design, affordable at scale, and culturally sensitive in ways that existing mHealth interventions are not.

The platform is not a chatbot, not a translation service, and not an AI medical advisor. It is a modular communication system that converts a clinician’s discharge decisions into actionable, voice-first, community-appropriate explanations — and then supports the patient through their recovery with proactive reminders, availability notifications, and on-demand information access, all on the device they already own.

If the pilot demonstrates feasibility and preliminary comprehension benefit, the implications extend well beyond oncology at Silchar Cancer Centre. Every regional hospital in India — and across the developing world — discharges patients with instructions they cannot fully understand, in formats they cannot easily use, in languages or registers that do not match their lived experience. Aakhyan offers a technically proven, ethically grounded, and economically viable platform to close that gap — not only at discharge, but across the continuum of post-discharge care.

## Data Availability

This manuscript describes a system architecture and planned trial protocol. No primary data were collected or analysed. All cited statistics are from published literature.

